# How Do Neurotransmitter Pathways Contribute to Neuroimaging Phenotypes?

**DOI:** 10.1101/2024.04.26.24305395

**Authors:** Amir Ebneabbasi, Mortaza Afshani, Arman Seyed-Ahmadi, Varun Warrier, Richard A.I. Bethlehem, Timothy Rittman

## Abstract

Neuroimaging could accurately reflect human behaviour in health and disease, but the mechanism by which image-derived phenotypes correspond to neurotransmitter systems remains uncertain. Prior studies have explored spatial correlations between neuroimaging phenotypes and positron emission tomography radiotracers. However, the influence of neurotransmitters goes beyond the receptors/transporters, influencing a wider array of intracellular components as pivotal parts of neurotransmitter pathways. Here, we used unsupervised learning to understand how the brain maps of healthy function (i.e., magnetoencephalography frequency-specific power) and abnormal structure (i.e., disorder-specific cortical thickness) are closely anchored to underlying neurotransmitter pathways assessed by gene expression data. To do this, we used large-scale datasets of the Human Connectome Project (HCP), Enhancing NeuroImaging Genetics through Meta-Analysis (ENIGMA) and Allen Human Brain Atlas (AHBA). We considered spatial and random gene null models to mitigate false positives. We replicate our analyses using different gene stability thresholds. This analytic approach paves the way for personalised medicine and advanced biomarkers.

## Introduction

The rapid advancement of technology has ushered in a new era of high-resolution neuroimaging, with methods including magnetoencephalography (MEG) and magnetic resonance imaging (MRI) providing unprecedented insights into neural oscillations and morphology (Bijsterbosch et al., 2017; Jenkinson & Chappell, 2018). These tools serve as proxies for understanding health and disease (Miller et al., 2016; Rittman, 2020). However, how these macroscopic measures arise from underlying molecular foundations remains unclear (Morgan et al., 2019; Seidlitz et al., 2020). Herein, neurotransmitters come to the forefront, playing a vital role in the tapestry of neurophysiology, even as our understanding of their specific contributions to neuroimaging phenotypes remains limited (Dukart et al., 2021; Goulas et al., 2021). These pathways contribute to orchestrating neural oscillations, underlining their indispensable role in routine brain functions (Baillet, 2017; Duncan et al., 2014). Moreover, dysregulations in the neurotransmitter system can significantly impact brain structure in psychiatric disorders (Sadock et al., 2015).

Previous research has begun associating image-derived phenotypes with neurotransmitter receptor maps obtained from positron emission tomography (PET) (Hänisch et al., 2023; Hansen et al., 2021; Hettwer et al., 2022). While PET receptors have provided valuable insights into the mapping of neurotransmitter systems within the brain, there are reasons why employing gene expression data from the Allen Human Brain Atlas (AHBA) (Hawrylycz et al., 2012) offers an alternative perspective to uncover the neurochemical underpinnings of neuroimaging profiles. First, neurotransmitter pathways intricately encompass synthesis, release, metabolism, and re-uptake mechanisms, constituting a complex network. From a pharmacological perspective, therapeutic targets extend to receptors and intracellular markers within these pathways (Hansen, Markello, et al., 2022; Luppi et al., 2023). AHBA covers a diverse spectrum of genes involved in neurotransmission, potentially providing a more comprehensive view of these intricate pathways. Second, the existing literature has often disregarded individual variability in the topographic distribution of neurotransmitters (Markello et al., 2022), focusing on general trends rather than personalised differences. By employing AHBA, the differential stability (DS) concept is introduced, and assessed as the similarity of transcriptional activity among six post-mortem donors (Hawrylycz et al., 2015). This concept paves the way for exploring individual differences in gene expression, allowing a more nuanced understanding of how specific neurotransmitter pathways contribute to distinct neuroimaging profiles. Third, using gene markers to define neurotransmitter pathways creates the potential for conducting genetic sensitivity analyses.

Our study pioneers the relationship between neurotransmitter pathways assessed via gene expression data from AHBA and neuroimaging profiles. We use large-scale datasets of the Human Connectome Project (HCP-MEG) (Van Essen et al., 2013) and Enhancing NeuroImaging Genetics through Meta-Analysis (ENIGMA) (Thompson et al., 2020) to specify whether the spatial patterns of healthy neural function (i.e., frequency-specific power maps) and abnormal structure (i.e., case-control cortical thickness maps) are co-located with neurotransmitter pathways. We leverage partial least squares for each neuroimaging map to rank the predictor weights of brain genes. Gene enrichment analysis (Subramanian et al., 2005) is then used to explore whether neurotransmitter-specific gene sets are positioned within the extremities of each PLS-ranked gene list. We incorporate both spatial and random gene null models to address the potential issue of false positives. We also replicate our findings using different DS thresholds (Hawrylycz et al., 2015). The insights gleaned from our study could have significant implications for personalised medicine and the advancement of biomarkers.

## Results

### Neurotransmitters: body tissues, brain pathways and neuropsychological functions

To study the neurochemical architecture of imaging phenotypes, we initially extracted the neurotransmitter-specific gene markers from AMIGO2 (https://amigo.geneontology.org/amigo) (Carbon et al., 2009). We meticulously curated sets of genes for each neurotransmitter, encompassing key aspects such as metabolism, transport, binding, and re-uptake (Supplementary Excel File). The included neurotransmitters are GABA, glutamate, dopamine, serotonin, histamine, aspartate, glycine, epinephrine, norepinephrine, and acetylcholine. The compiled gene sets for these ten neurotransmitters have been outlined in the Supplementary Excel File and Figure 1a.

**Figure 1.**
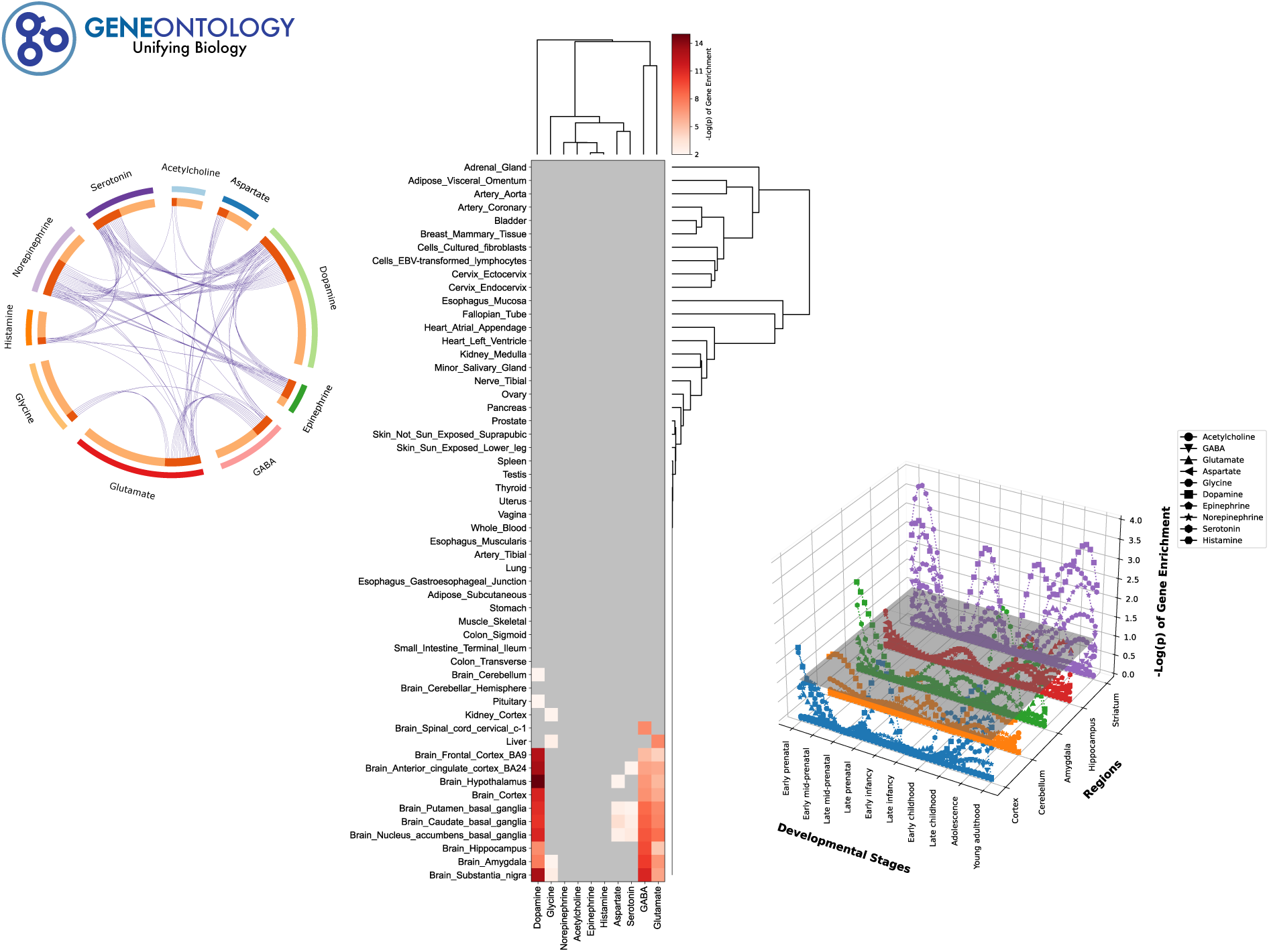
(a) Ontology, (b) developmental profile and (c) cross-tissue expression of neurotransmitter-specific genes. (a) To study the neurochemical architecture of imaging phenotypes, we initially extracted the neurotransmitter-specific gene markers from AMIGO2. We curated sets of genes for each neurotransmitter, encompassing key aspects such as metabolism, transport, binding, and re-uptake. (b) The Genotype-Tissue Expression (GTEx) database was employed to assess the extent of overlap between gene sets specific to neurotransmitters and those of body tissues. Log10 FDR-corrected p-values of < 0.05 are colorised. (c) BrainSpan data was utilised to discern the potential contribution of neurotransmitters to developmental stages. Log10 FDR-corrected p-values of < 0.05 are represented above the grey plane.

To gain insights into the potential role of neurotransmitters across diverse body tissues, we utilised the Genotype-Tissue Expression (GTEx) dataset (https://gtexportal.org/home/) (Keen & Moore, 2015). We assessed whether gene sets specific to neurotransmitters show disproportionate overlap with differentially expressed genes (DEG) in tissues. For this purpose, we employed hypergeometric tests facilitated by the FUMA tool available at (https://fuma.ctglab.nl/) (Watanabe et al., 2017). The False Discovery Rate (FDR) (Benjamini & Hochberg, 1995) was applied to take multiple comparisons into account. Our investigation revealed a significant upregulation of GABA, glutamate, dopamine, serotonin, aspartate, and glycine within the brain compared to other body tissues (Figure 1b). As a subsequent step, we examined potential intersections between gene sets linked to neurotransmitters and those derived from different developmental profiles sourced from the BrainSpan dataset (http://www.brainspan.org) (Miller et al., 2014). We used the CSEA tool available at (http://genetics.wustl.edu/jdlab/csea-tool-2/) to perform the FDR-corrected enrichment analysis. Our results revealed that dopamine and serotonin exhibited more pronounced transcriptional alterations across developmental stages, with a notable emphasis within the striatum, as depicted in Figure 1c.

To trace the expression pattern of neurotransmitter pathways throughout the whole brain, we employed the Allen Human Brain Atlas (AHBA) (https://human.brain-map.org/) (Hawrylycz et al., 2012) while adhering to the recent best-practice guidelines for its data preprocessing (Larivière et al., 2021; Markello et al., 2021). This remarkable resource encompasses an extensive RNA microarray survey covering “all genes, all structures” in six post-mortem brains. These gene expression profiles are aligned with the standardised space of the brain (MNI), enabling precise spatial localisation. We assigned the transcriptional activity of 12,668 genes to the Desikan-Killiany atlas (DK) (Desikan et al., 2006). Given that only two of the six donors possessed data for the right hemisphere, we focused our analysis on the left hemisphere. To visualise brain patterns, we calculated the average transcriptional values within each neurotransmitter gene set (Supplementary Figure 1).

As a preliminary step toward translating molecular pathways into neuropsychological patterns, we conducted Spearman correlation analyses between neurotransmitter maps and term-based meta-analytic-driven spatial phenotypes sourced from the Neurosynth database (https://neurosynth.org/) (Yarkoni et al., 2011). We examined 123 terms from the Cognitive Atlas (Poldrack et al., 2011), a publicly available cognitive science ontology. These terms have been previously employed alongside the Neurosynth (Alexander-Bloch et al., 2018; Hansen et al., 2021). The selected terms encompass a spectrum, ranging from overarching concepts (such as attention and emotion) to more specific cognitive processes (such as visual attention and episodic memory), behaviours (including eating and sleep), and emotional states (like fear and anxiety). The meta-analytic-driven probability maps provided by Neurosynth were then parcellated into the DK atlas. Results are shown with word clouds in supplementary Figure 2. Many neurotransmitters are associated with affective functions. Among them, epinephrine and glycine are notably associated with cognitive terms.

### Neurotransmitters and healthy brain oscillations

To interrogate the cortical source of diverse neural oscillations, we employed group-level source-localised power maps of magnetoencephalography (MEG) records within a cohort of 33 genetically unrelated healthy participants. These power maps were accessed through the *neuromaps* (https://netneurolab.github.io/neuromaps/usage.html) (Markello et al., 2022; Shafiei et al., 2022), which in turn were derived from the Human Connectome Project (HCP) (https://www.humanconnectome.org/study/hcp-young-adult) (Elam et al., 2021; Glasser et al., 2013). Six power maps were included as follows: delta (2–4 Hz), theta (5–7 Hz), alpha (8–12 Hz), beta (15–29 Hz), low gamma (30–59 Hz) and high gamma (60–90 Hz) (Figure 3).

To compute the transcriptional foundation of each power map, a partial least squares (PLS) analysis [17] was conducted. PLS aims to find components from the gene expression matrix (34 regions × 12,668 genes) that have maximal covariance with each power map (34 regions × 1). We applied 10,000 spin permutations to account for the spatial autocorrelation of brain regions (Alexander-Bloch et al., 2018). FDR correction (Benjamini & Hochberg, 1995) was also performed to compensate for the multiple PLS tests conducted (i.e., six times). PLS1 weighted maps were significantly correlated with power maps (*p _spin-FDR_ values were all <* 0.01) (r _delta_ = 0.81, *R^2^* = 0.65; r _theta_ = 0.72, *R^2^*= 0.52; r _alpha_ = 0.81, *R^2^* = 0.65; r _beta_ = 0.83, *R^2^* = 0.68; r _low-gamma_ = 0.77, *R^2^*= 0.60; r _high-gamma_ = 0.78, *R^2^* = 0.60). Explained variance (*R^2^*) and p-value of other PLS components are provided in the Supplementary Excel File. We did not include the PLS2 component in subsequent analyses since it was not significant in all power maps and hindered us from comparing the transcriptional foundation of different power maps (Supplementary Figure 3). Furthermore, to estimate the reliability of the PLS1-driven gene weights, we randomly rearranged the rows (brain regions) of the gene expression matrix and repeated the PLS analysis. This procedure was conducted 10,000 times to create a null distribution for each gene. The original weight of each gene was then divided by the standard error of its corresponding null distribution (Morgan et al., 2019). Genes with higher bootstrapped weights significantly contribute to PLS1. The bootstrapped gene weights of PLS1 can be found in the Supplementary Excel File.

To explore whether neurotransmitter gene sets are positioned within the extremities of each PLS1-ranked gene list, we leveraged the fast gene set enrichment analysis (FGSEA) (https://www.gsea-msigdb.org/gsea/index.jsp) (Subramanian et al., 2005). Enrichment scores of neurotransmitters were corrected (Benjamini & Hochberg, 1995) and normalised to manage the number of gene sets and variations in their sizes, respectively. Agglomerative hierarchical clustering was also employed to gauge the similarity of power maps based on normalised enrichment scores (NES). The methodological overview of the current study is depicted in Figure 2. It is worth noting that FGSEA offers clear advantages over the hypergeometric approach by considering the complete range of PLS-driven gene rankings. This eliminates the necessity of arbitrary thresholds to define outlier bounds and employs a robust permutation technique involving ’random genes’ to estimate the statistical significance (Subramanian et al., 2005). The implication of selecting either absolute or signed PLS-driven genes in the context of enrichment analysis has been discussed previously (Romero-Garcia et al., 2019). According to the overall pattern of FGSEA findings (Figure 3 and Supplementary Figure 4), the power maps were positively associated with the neurotransmitter pathways, except for alpha. In addition, maps were mostly shaped by the histaminergic pathway.

**Figure 2.**
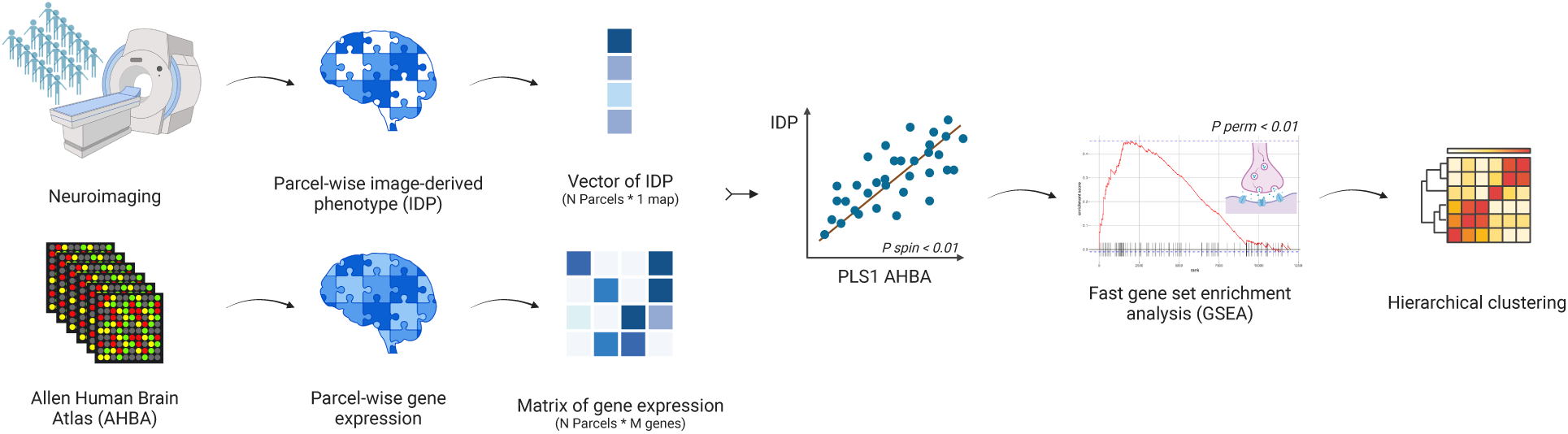
Overview of imaging-transcriptomic co-location, followed by neurotransmitter enrichment analysis. We used large-scale neuroimaging datasets to specify whether the spatial patterns of health and disease are co-located with neurotransmitter pathways. We leveraged partial least squares (PLS) for each neuroimaging map to rank the predictor weights of AHBA genes. Gene enrichment analysis was used to explore whether neurotransmitter-specific gene sets are positioned within the extremities of each PLS-ranked gene list. Hierarchical clustering was also employed to gauge the similarity of imaging maps based on neurotransmitter enrichment scores. The figure was created by BioRender.

**Figure 3.**
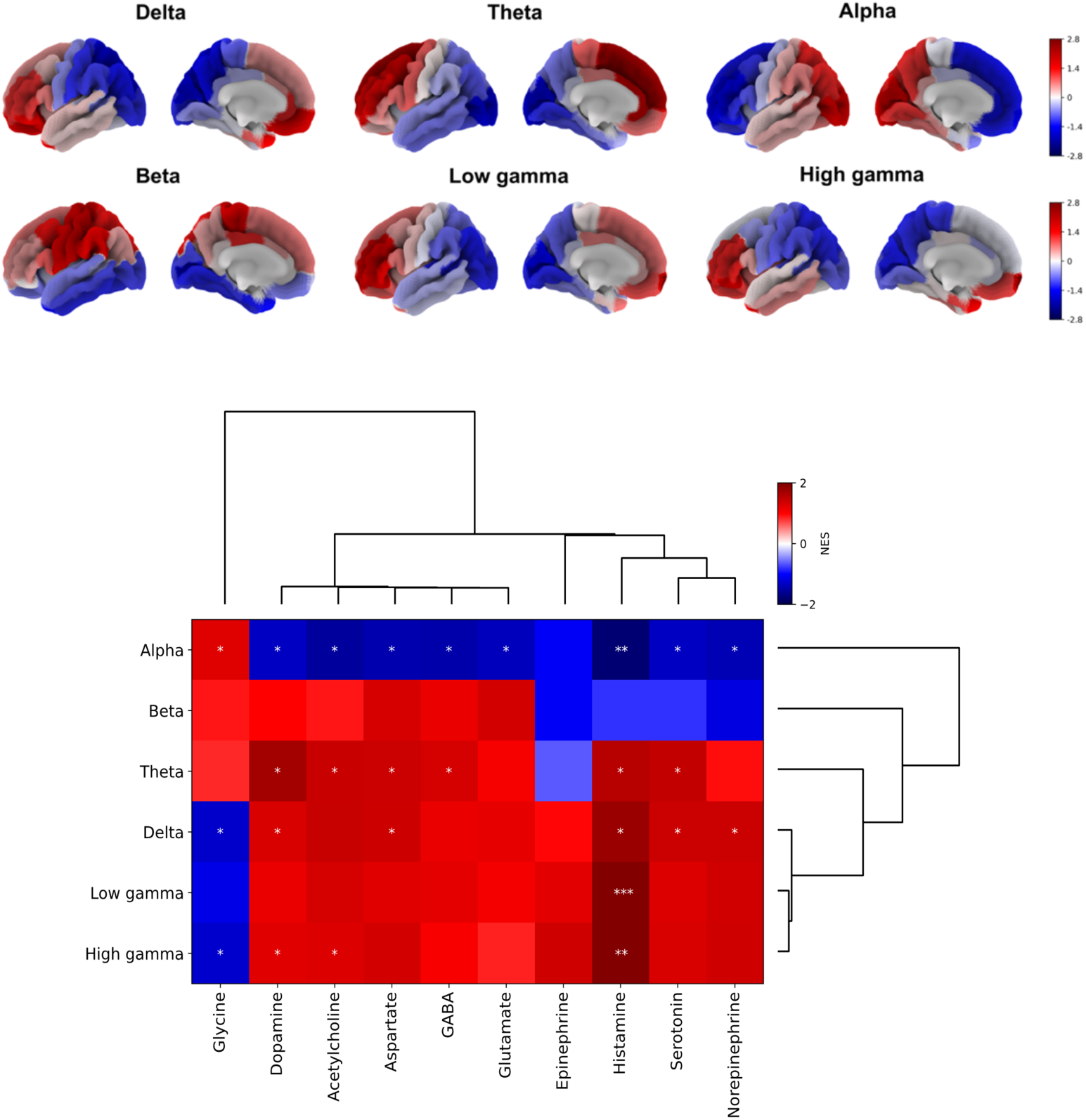
Power maps of frequency bands in Magnetoencephalography, and their taxonomy according to normalised enrichment scores (NES) of neurotransmitter-specific genes. (a) The Human Connectome Project provides resting-state MEG signals that can be used to generate group-level source-localised power maps. Six frequency-specific maps were created for delta (2-4 Hz), theta (5-7 Hz), alpha (8-12 Hz), beta (15-29 Hz), low gamma (30-59 Hz), and high gamma (60-90 Hz) bands. Higher power is denoted by positive values represented in red. (b) Whole-brain expression of genes was taken from the Allen Human Brain Atlas. To rank the predictor weights of 12,668 genes, a partial least squares (PLS) analysis with brain region bootstrapping and spin permutation testing was performed for each power map. We utilised fast gene set enrichment analysis (FGSEA) to investigate whether neurotransmitter gene sets are positioned within the extremities of each PLS-ranked gene list with a standard FDR-corrected p-value cutoff of < 0.25 to account for normalised enrichment scores (NES). P-values are marked with one (< 0.25), two (< 0.05) and three (< 0.01) asterisks. Finally, we applied hierarchical clustering to assess the similarity of power maps based on the NES of neurotransmitters.

### Neurotransmitters and abnormal brain structure

To interrogate the neural foundation of psychiatric disorders, we obtained covariate-adjusted case-control cortical thickness (ΔCT) maps from the *ENIGMA toolbox* (https://enigma-toolbox.readthedocs.io/en/latest/) (Larivière et al., 2021). The ENIGMA consortium adopts standard quality control and pre-processing protocols (Thompson et al., 2020). We included cohorts of autism spectrum disorder (ASD; 1571 patients, 1651 controls) (van Rooij et al., 2018), attention-deficit/hyperactivity disorder (ADHD; 733 patients, 539 controls) (Hoogman et al., 2019), schizophrenia (SCZ; 4474 patients, 5098 controls) (van Erp et al., 2018), bipolar disorder (BD; 1837 patients, 2582 controls) (Hibar et al., 2018), major depressive disorder (MDD; 2148 patients, 7957 controls) (Schmaal et al., 2017) and obsessive-compulsive disorder (OCD; 1905 patients, 1760 controls) (Boedhoe et al., 2018) (Figure 4).

**Figure 4.**
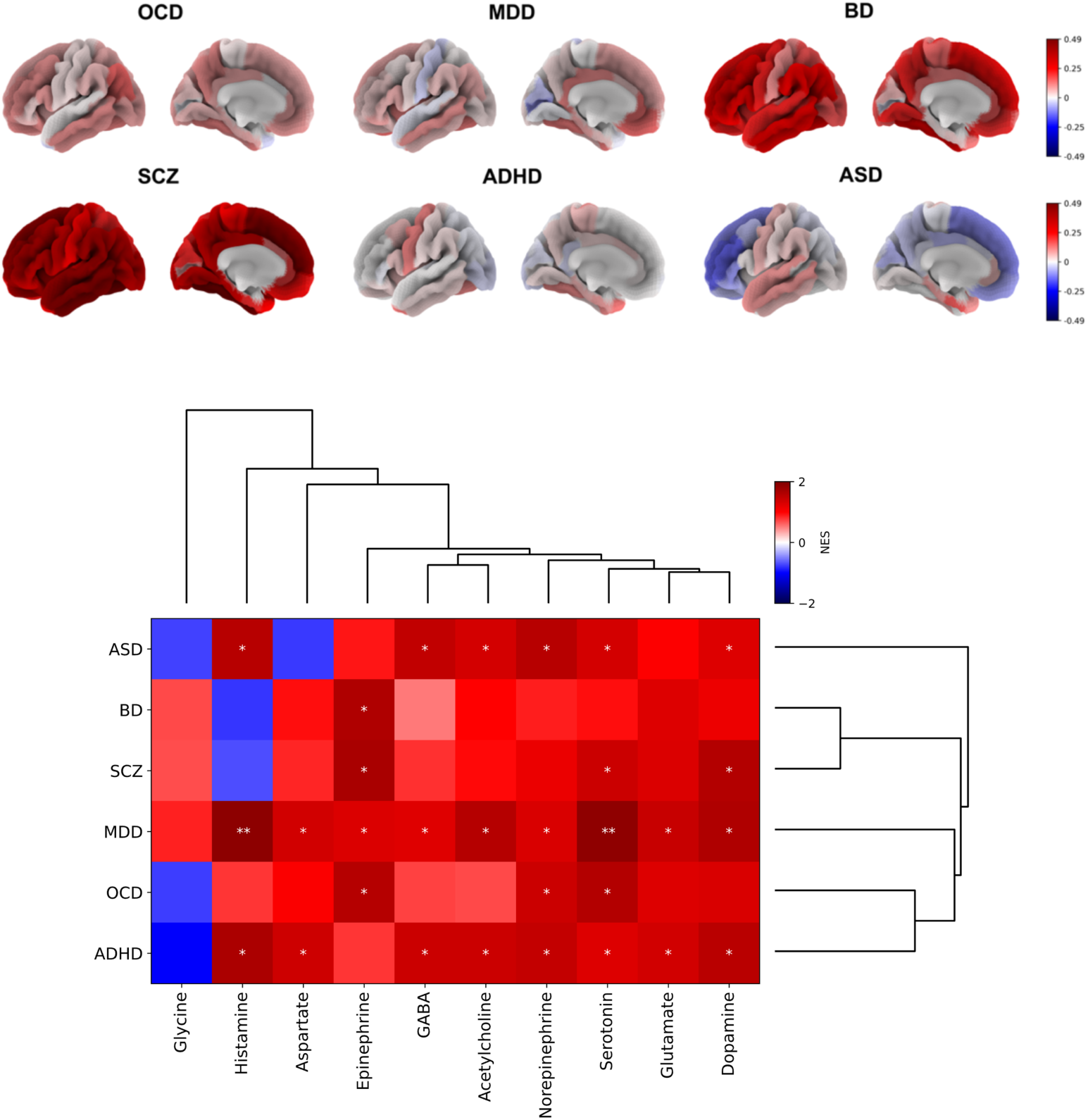
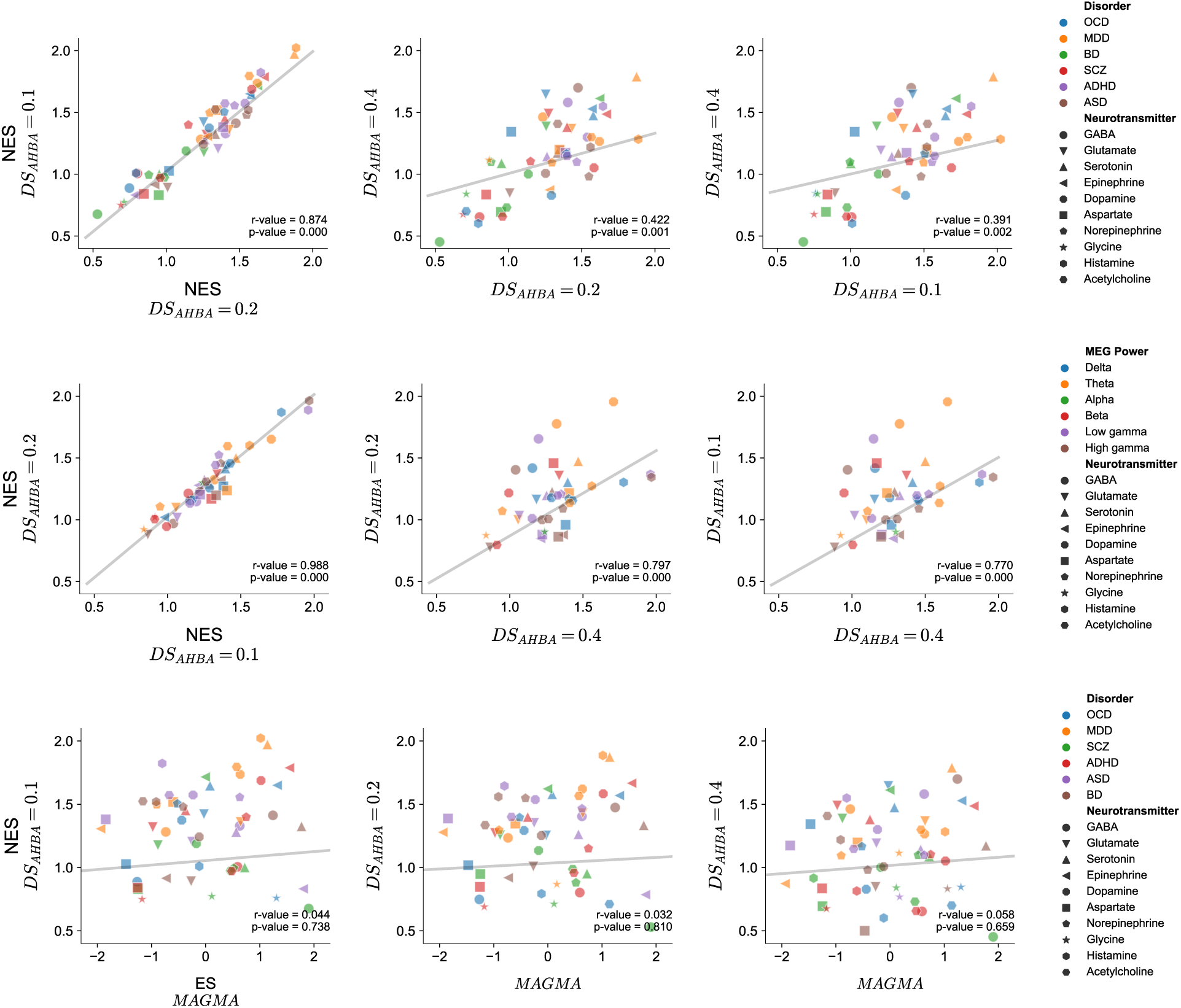
Case-control cortical thickness (CT) maps, and their taxonomy according to normalised enrichment scores (NES) of neurotransmitter-specific genes. (a) The ENIGMA consortium provided case-control maps of cortical thickness differences for attention-deficit/hyperactivity disorder (ADHD), autism spectrum disorder (ASD), schizophrenia (SCZ), bipolar disorder (BD), major depressive disorder (MDD), and obsessive-compulsive disorder (OCD). The raw data is pre-processed according to standard ENIGMA quality control protocols. Positive values (represented by red colors) indicated cortical thinning (i.e., shrinkage) in patients compared to controls. (b) PLS, FGSEA and hierarchical clustering were repeated for cortical thickness (CT) maps. Gene-imaging spatial co-location was explored using partial least squares (PLS) analysis. Fast gene set enrichment analysis (FGSEA) was then applied using neurotransmitter-specific gene sets. To ensure the robustness of our findings, we conducted PLS and FGSEA analyses while employing varying DS thresholds. We explored both lenient (r > 0.1, involving 15,631 genes) and more stringent thresholds (r > 0.4, encompassing 6,513 genes), in comparison to the original dataset, which employed a threshold of r > 0.2 and 12,668 genes. This analysis consistently revealed enrichment of neurotransmitter-related pathways across the spectrum of DS thresholds. In our sensitivity analysis, we examined potential overlaps between gene sets specific to neurotransmitters and alternative psychiatric gene candidates. Pearson correlations did not reveal a consistent enrichment pattern across the PLS- and GWAS-derived gene candidates. Nonetheless, we replicated the significance of serotonin (Note: negative values of NES and ES are not shown in the plots for better visualisation).

PLS analysis was repeated for each ΔCT map. PLS1 weighted maps were significantly correlated with ΔCT maps (*p _spin-FDR_ values were all <* 0.01) (r _ASD_ = 0.60, *R^2^* = 0.37; r _ADHD_ = 0.62, *R^2^* = 0.39; r _SCZ_ = 0.71, *R^2^* = 0.50; r _BD_ = 0.78, *R^2^* = 0.60; r _MDD_ = 0.66, *R^2^* = 0.44; r _OCD_ = 0.70, *R^2^* = 0.49). *R^2^* and p-value of other PLS components are provided in the Supplementary Excel File. We did not include the PLS2 component in subsequent analyses since it was not significant in all ΔCT maps (Supplementary Figure 5). Bootstrapped PLS1-driven gene weights are included in the Supplementary Excel File. According to the overall pattern of FGSEA findings (Figure 4 and Supplementary Figure 6), ΔCT maps were positively co-located with the neurotransmitter pathways. We found that some disorders (MDD, ADHD and ASD) are more widely associated with neurotransmitter pathways than others (BD, SCZ and OCD). We also observed that most ΔCT maps are anchored with the serotonergic pathway. In addition, we found dopamine to be significantly associated with SCZ, ADHD, MDD and ASD. On the other hand, we found relationships with some support in the literature but, to our knowledge, have not been convincingly translated into clinical disorders. For instance, our findings suggested a potential role of histamine in MDD, ADHD, and ASD, along with the involvement of epinephrine in SCZ and BD.

### Robustness and sensitivity analysis

To address the robustness of the findings, PLS and FGSEA were repeated using a range of differential stability (DS) thresholds. This approach allowed us to consider individual differences in gene expression, leading to a more nuanced understanding of how robust neurotransmitter pathways contribute to neuroimaging phenotypes. There is evidence that genes with higher DS exhibit lower interpersonal variability and stronger biological relevance (Hawrylycz et al., 2015). We considered both lenient (r > 0.1, n = 15631 genes) and stricter DS thresholds (r > 0.4, n = 6513 genes) in comparison to the original dataset (r > 0.2, n = 12,668 genes). Pearson correlations showed a convergent pattern of NES across the three DS thresholds (Figure 5).

For sensitivity analysis, we delved into the link between inherited genetic risk and neurotransmitter abnormalities in psychiatric disorders using GWAS summary statistics from the Psychiatric Genomic Consortium (https://pgc.unc.edu/for-researchers/download-results/) and applying MAGMA (multi-marker analysis of GenoMic annotation) (https://ctg.cncr.nl/software/magma) (de Leeuw et al., 2015). Initially, we mapped the GWAS-driven SNPs to specific gene locations. SNPs were selected for six psychiatric disorders: ASD (18,381 patients, 27,969 controls) (Grove et al., 2019), ADHD (38,691 patients, 186,843 controls) (Demontis et al., 2023), SCZ (76,755 patients, 243,649 controls) (Trubetskoy et al., 2022), BD (41,917 patients, 371,549 controls) (Mullins et al., 2021), MDD (43,204 patients, 95,680 controls) (Wray et al., 2018) and OCD (2688 patients, 7037 controls) (OCGAS, 2018). Gene boundaries were defined using Ensembl release 92 (GRCh37) and extended by 35 kilobase pairs (kb) upstream and 10 kb downstream to encompass potential regulatory regions. Next, we converted the SNP p-values into gene-level metrics. The European panel of 1,000 Genomes phase 3 was utilised to account for linkage disequilibrium (LD). Competitive gene-set analysis was then conducted. The neurotransmitter-specific p-value assesses whether genes within the gene set exhibit a higher average association with the phenotype of interest (here, psychiatric disorders) than genes outside the set. P-values of *βs* were corrected for multiple comparisons (Benjamini & Hochberg, 1995). Pearson correlations did not reveal a consistent enrichment pattern across the PLS- and GWAS-derived gene candidates (Figure 5). Nonetheless, we replicated the significance of serotonin, with its p-value ranking first in BD (P< 0.02) and (P< 0.15) MDD and second in SCZ (P< 0.07).

## Discussion

Image-derived phenotypes could reflect human behaviour in health and disease, though the translation of these phenotypes into neurophysiology remains incompletely understood. While previous research has explored the co-location of neuroimaging markers with PET-derived receptors/transporters (Hänisch et al., 2023; Hansen et al., 2021), the Allen Human Brain Atlas (AHBA) (Hawrylycz et al., 2012) emerges as a promising alternative. First, AHBA’s expansive coverage of genes linked to intracellular components widens the lens beyond prevailing receptor-focused investigations. Second, AHBA introduces the concept of differential stability (DS) to address the often overlooked individual variability in the neurotransmitter distribution (Hawrylycz et al., 2015). Third, using gene markers to delineate neurotransmitter pathways opens the potential for conducting gene-focused sensitivity analyses.

In this study, we investigated how the brain maps of healthy function (i.e., frequency-specific power maps of HCP-MEG (Van Essen et al., 2013)) and abnormal structure (i.e., case-control cortical thickness (ι1CT) maps of ENIGMA (Thompson et al., 2020)) are closely anchored to underlying neurotransmitter pathways assessed using the AHBA (Hawrylycz et al., 2012). We selected AMIGO2 (Carbon et al., 2009) to extract gene markers associated with various dimensions of each neurotransmitter’s operation, encompassing metabolism, transport, binding, and re-uptake. Partial least squares (PLS) identified components from the gene expression matrix with maximal covariance with each imaging map. To address the spatial autocorrelation of brain regions, we applied spin permutation. Fast gene set enrichment analysis (FGSEA) was employed to determine the positioning of neurotransmitter gene sets within the PLS-ranked gene lists. We accounted for the difference in gene set sizes and the number of gene sets.

We showed that neural correlates of healthy function are positively associated with different neurotransmitter pathways. A recent study also found that canonical maps of MEG are shaped by the overlapping spatial patterns of multiple PET receptors (Hansen, Shafiei, et al., 2022). This observation aligns with Pharmaco-MEG evidence that a wide range of pharmacological substances affects the MEG signal (Duncan et al., 2014; Gross, 2019). How those neurotransmitters interact to induce complex brain functions reflected in different frequency-specific signal components remains unclear. In addition, we observed that power maps exhibited a prefrontal gradient of power (delta, theta, low- and high-gamma) mostly linked to the histaminergic pathway. We replicated previous findings (Hansen, Shafiei, et al., 2022) that the spatial distribution of histamine radiotracer (H3) significantly contributes to the power maps. Another study observed that histamine amalgamates local theta and gamma power in MEG (Chen et al., 2017). Histaminergic neurons in the brain serve multiple signalling pathways. As a key regulator of whole-brain activity, histamine could modulate the release of other neurotransmitters (Shahid et al., 2011; Wada et al., 1991). Activation of a small group of tuberomammillary nucleus cells in the hypothalamus results in the release of histamine, which subsequently upregulates neural excitability in widespread target cells across the brain (Haas & Panula, 2003).

We also indicated that ι1CT maps across neuropsychiatric conditions are positively co-located with different neurotransmitter pathways. This suggests that the heterogeneity of symptoms in the clinical profile of psychiatric patients might be partly related to the various types of neurotransmitters involved. Put differently, disorders are not discrete islands of abnormalities but continuums of interconnected phenomena that are quantitatively differentiated (Khodadadifar et al., 2022; Linscott & van Os, 2010). Several works have delineated the transdiagnostic nature of cortical abnormalities (Hettwer et al., 2022; Patel et al., 2021). A recent study discovered a shared dimension of cortical anomaly across psychiatric disorders linked to microstructure gradients and serotonin and dopamine receptor distributions (Park et al., 2022). Consistent with these findings, our study demonstrated a significant association between the serotonergic pathway and ΔCT maps, providing further evidence for the transdiagnostic explanation of mood dysfunction (Husain & Roiser, 2018). In addition, dopamine was significantly engaged in SCZ, ADHD, MDD and ASD, highlighting its widespread role in cognition, affect and locomotion (Grace, 2016; Ott & Nieder, 2019).

To address the robustness of our findings, we repeated the PLS and FGSEA analyses by applying varying DS thresholds. We considered both lenient (r > 0.1, n = 15631 genes) and stricter DS thresholds (r > 0.4, n = 6513 genes) in comparison to the original dataset (r > 0.2, n = 12,668 genes). We uncovered a consistent trend of neurotransmitter enrichment across the range of DS thresholds. This demonstrates the robustness of the interplay between neurotransmitters and neuroimaging phenotypes, even in the presence of individual differences in gene expression. In the sensitivity analysis, we investigated potential overlaps between neurotransmitter-specific genes and psychiatric gene candidates with an inherited component. Pearson correlations did not reveal a uniform enrichment pattern among the PLS- and GWAS-derived gene candidates. Nonetheless, we confirmed the significance of serotonin in mood disorders. It is important to note that neurotransmitter dysfunctions may be linked to post-translational processes and are not solely inherited (Tsankova et al., 2007; Yuan et al., 2023).

This study is not without limitations. First, transcriptional activity and in-vivo density of some neurotransmitters are spatially correlated (e.g., serotonin and dopamine), and that of others is debated (Hansen, Markello, et al., 2022). The poor topographic correspondence is partly related to genes with higher subject variability (i.e., lower DS). Nonetheless, we replicated our findings using a range of DS thresholds. The second limitation points to the brain segregation assumption of the gene-imaging analysis. This could oversimplify the interconnected nature of brain regions. Neurotransmitters acting within one brain region affect distant areas through structural connectivity. Future investigations could benefit from adopting a multimodal perspective that captures network dynamics and interactions. Thirdly, the spatial co-location of neurotransmitter pathways and neural correlates of disease do not necessarily indicate causal relationships. To overcome this limitation would require causal methodologies, such as Mendelian Randomisation (Sanderson et al., 2022; Taschler et al., 2022). This approach would enable an exploration of whether single nucleotide polymorphisms (SNPs) predicting neurotransmitter changes also affect the neural correlates of disease (Stauffer et al., 2023; Warrier et al., 2021).

In conclusion, we considered an imaging-transcriptomic analysis to explore the neurochemical foundation of image-derived phenotypes in health and disease. We showed that various neurotransmitters contribute to neural oscillations, with a central role for the histaminergic pathway. Furthermore, we revealed an association between psychiatric disorders and a wide array of neurotransmitter pathways, with a particular emphasis on the serotoninergic pathway. Our methodology encompassed spatial and random gene null models to mitigate false positives. We replicated our findings using different gene stability thresholds. Future investigations could deepen our understanding by integrating genetic and epigenetic factors, along with causal inference, to provide a more nuanced comprehension of the intricate neurochemical processes within the brain.

## Methods

### Frequency-specific power

MEG data was obtained from the Human Connectome Project (1200 release). Acquisition protocols and pre-processing of resting-state MEG are provided elsewhere (Elam et al., 2021; Glasser et al., 2013). MEG power analysis was performed in a cohort of 33 unrelated participants (age range: 22–35 years, 17 males) (Markello et al., 2022; Shafiei et al., 2022). In brief, Brainstorm’s linearly constrained minimum variance (LCMV) beamformers method (Tadel et al., 2011) was used to extract the source activity for each participant in six frequency bands: delta (2–4 Hz), theta (5–7 Hz), alpha (8–12 Hz), beta (15–29 Hz), low gamma (30–59 Hz) and high gamma (60–90 Hz). Welch’s method was leveraged to estimate power spectrum density (PSD) for the source-level data. The average power at each frequency band was then calculated for the DK atlas.

### Case-control cortical thickness (ΔCT)

The ENIGMA (Enhancing Neuroimaging Genetics through Meta-Analysis) consortium is a data-sharing initiative that follows standardised image acquisition and processing pipelines for exploring the genetic and neural foundations of brain diseases (Thompson et al., 2020). The ENIGMA toolbox (Larivière et al., 2021) was used to obtain covariate-adjusted case-vs-control cortical maps. These parcel-wise values were determined by conducting cross-site random-effect meta-analyses on Cohen’s d-values calculated for cortical thickness. Multiple linear regression analyses were performed to account for age, sex, and site. For more information on the imaging and quality control protocols, see http://enigma.ini.usc.edu/protocols/imaging-protocols. In our study, case-vs-control effect sizes were collected for six psychiatric disorders: attention-deficit/hyperactivity disorder (ADHD; 733 patients, 539 controls), autism spectrum disorder (ASD; 1571 patients, 1651 controls), schizophrenia (SCZ; 4474 patients, 5098 controls), bipolar disorder (BD; 1837 patients, 2582 controls), major depressive disorder (MDD; 2148 patients, 7957 controls) and obsessive-compulsive disorder (OCD; 1905 patients, 1760 controls).

### Neuropsychological translation

Neurosynth is an online platform for the automated synthesis of functional magnetic resonance imaging (fMRI) experiments (Yarkoni et al., 2011). Firstly, text-mining techniques are employed to identify neuroimaging studies utilising specific terms of interest at a high frequency (>1 in 1,000 words). Secondly, peak coordinates are extracted from the reported tables. This approach results in a large-scale database of term-to-coordinate mappings. Thirdly, whole-brain meta-analyses are conducted to quantify brain-cognition relationships. Neurosynth results should be interpreted with caution. It fails to differentiate between activated or deactivated areas and disregards the degree of activation. Indeed, it simply highlights that certain brain areas are frequently mentioned with certain words. We examined 123 terms from the Cognitive Atlas (Poldrack et al., 2011), a publicly available cognitive science ontology.

### Transcriptomic data

This study utilised regional microarray expression data from postmortem brains sourced from the Allen Human Brain Atlas (AHBA) (Hawrylycz et al., 2012). The analysis focused on the left hemisphere due to limited samples from the right hemisphere. Data was processed using the abagen toolbox (Markello et al., 2021). Several preprocessing steps were employed: probe-to-gene re-annotation, probe intensity-based filtering, probe selection, sample-to-region assignment, sample normalisation, gene normalisation, and sample-to-region combination.

First, microarray probes underwent reannotation (Arnatkevičiūtė et al., 2019). Probes that could not be linked to a valid Entrez ID were excluded from the analysis. Following that, a filtration process was implemented for the probes. Probes exhibiting intensity levels lower than the background noise in ≥ 50% of the samples across donors were excluded from the dataset. In cases where multiple probes indexed the expression of the same gene, we selected the probe that displayed the most consistent pattern of regional variation across donors. This was determined by the level of differential stability (DS) (Arnatkevičiūtė et al., 2019), which was computed using the following formula:

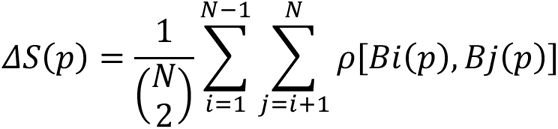

Let *ρ* represent Spearman’s rank correlation of the expression of a single probe, denoted as ’p’, across regions in two donor individuals, Bi and Bj. N stands for the total number of donors. In this context, the term “regions” pertains to the structural designations outlined in the ontology of AHBA.

Next, The MNI coordinates of tissue samples were revised to align with those obtained through non-linear registration using Advanced Normalization Tools (https://github.com/chrisgorgo/alleninf). Samples were allocated to specific brain regions by minimising the Euclidean distance between the MNI coordinates of each sample and the closest surface vertex. Samples for which the Euclidean distance to the nearest vertex exceeded two standard deviations above the mean distance for all samples from the same donor were excluded. In cases where a brain region lacked sample assignments from any donor through the procedure, the tissue sample closest to the centroid of that parcel was independently identified for each donor. The average expression values of these identified samples were computed across donors, with weights assigned based on the distance between the parcel centroid and each sample. This provided an estimate of the parcellated expression values for the missing region. All tissue samples that couldn’t be assigned to a brain region as defined in the provided atlas were discarded. To account for inter-subject variation, expression values were normalised across genes using a robust sigmoid function:

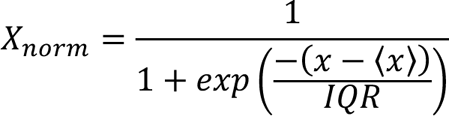

Where ⟨x⟩ represents the median, and IQR signifies the normalised interquartile range of the expression of a single tissue sample across genes. Subsequently, the normalised expression values were rescaled to fit within the unit interval:

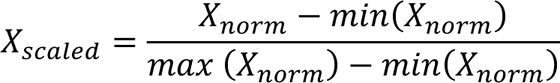

Gene expression values were subsequently normalised across tissue samples using the same procedure. Samples assigned to the same brain region were averaged independently for each donor and then aggregated across all donors, resulting in a regional expression matrix. However, due to the limited number of samples available in the right hemisphere (data were only available for two donors), regions in the right hemisphere were excluded. Finally, we obtained expression values for 12,668 unique genes across 34 Desikan-Killiany atlas (DK) regions (Desikan et al., 2006).

### Unsupervised learning

Partial least squares regression (PLS) is a widely recognised multivariate approach to identify links between a group of predictors and response variables. PLS can be applied when the number of predictor variables exceeds the number of observations or the predictors are not independent (Morgan et al., 2019; Vértes et al., 2016). PLS was used to find components from the gene matrix that have maximal covariance with each brain map. The normalised gene expression matrix (34 regions × 12,668 genes) was considered the X matrix, and the normalised imaging map (34 regions × 1) was selected as the Y matrix. The first PLS component (PLS1) was then selected for subsequent analyses. To estimate the reliability of the PLS1-driven gene weights, we randomly rearranged the rows (brain regions) of the gene expression matrix and repeated the PLS analysis. This procedure was conducted 10,000 times to create a null distribution for each gene. The original weight of each gene was then divided by the standard error of its corresponding null distribution (Morgan et al., 2019). Genes with higher bootstrapped weights significantly contribute to PLS1.

### Spatial null model

Spin permutation tests were used to confirm whether co-locations between PLS1 and image-derived phenotypes were irrelevant to the inherent autocorrelation of brain regions. We created null models representing the overlap between the X and Y maps. We achieved this by projecting the spatial coordinates of cortical maps onto surface spheres, applying randomly sampled rotations, and reallocating cortical values accordingly. Subsequently, we compared the original correlation coefficients with the empirical distribution of coefficients resulting from the spatial permutations (Alexander-Bloch et al., 2018; Markello & Misic, 2021).

### Gene set enrichment analysis

Inputs for Fast gene set enrichment analysis (FGSEA) (Subramanian et al., 2005):

1. Expression Data Set (D): The expression data set D comprises N genes.
2. Gene List (L): The L is generated through a ranking procedure alongside a phenotype or profile of interest, denoted as C. For each imaging-derived phenotype, we used PLS1-driven gene weights as L.
3. Exponent (p): This value is employed to control the weight of the step.
4. Gene Set (S): The S is independently derived and contains N_H_ genes, such as GO categories. We used AMIO2 to curate gene sets linked with various neurotransmitters, encompassing crucial functions like metabolism, transport, binding, and re-uptake. The neurotransmitters include GABA, glutamate, dopamine, serotonin, histamine, aspartate, glycine, epinephrine, norepinephrine, and acetylcholine.

Enrichment Score ES (S):

1. Rank the genes in set D, denoted as {g_1_, g_2_, …, g_N_}, based on the correlation, r (g_j_), between their expression profiles and C.
2. Calculate the proportion of genes in set S (’’hits’’) weighted by their correlation, and the proportion of genes not in set S (’’misses’’) present up to a specified position i in set L.

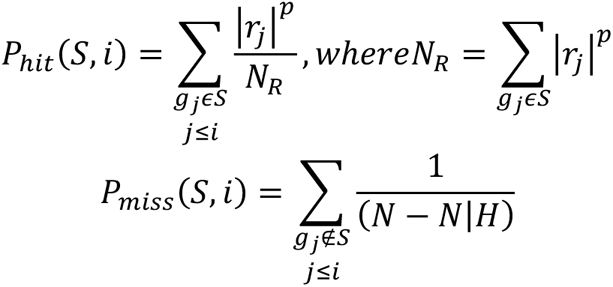 The Enrichment Score (ES) is the maximum deviation from zero, calculated as the difference between P_hit_ and P_miss_. In the case of a randomly distributed set S, ES (S) tends to be relatively small. However, if S is settled at the extremities of the gene list or exhibits a non-random distribution, ES (S) will correspondingly be high. When the parameter p is 0, ES (S) simplifies to the standard Kolmogorov–Smirnov statistic. When p is set to 1, the genes in set S are weighted based on their correlation with C, which is then normalised by the sum of correlations across all genes in S.

Estimating Significance:

1. A size-matched S is randomly selected with replacement, and the ES (S) is re-calculated.
2. The initial step is iterated 1,000 times through permutations, resulting in a distribution of ES _NULL_.
3. The nominal p-value of the original ES is calculated from the bimodal distribution of ES _NULL_.

Normalisation and p-value correction:

1. Adjust for variations in gene set size by normalising the observed ES (S). This step involves independently rescaling positive and negative scores, dividing them by the mean of the ES _NULL_ to generate the NES (S).
2. Calculate the False Discovery Rate (FDR). This step involves controlling the ratio of false positives to the total number of gene sets reaching a specified significance level. This is performed to control separately for positive and negative NES (S).

### Hierarchical clustering

We leveraged agglomerative hierarchical clustering to calculate the similarity of brain maps based on the NES of neurotransmitter-specific genes. First, a collection of n samples is arranged in m clusters, with m being equal to n. The method involves joining the most similar samples together to form a cluster, which leads to a decrease in the number of clusters by one after each iteration. This process continues until there is only one cluster left. The Euclidean distance technique assessed the distance between clusters, and the closest pair of subsets was identified using the Nearest Point Algorithm.

### BrainSpan

BrainSpan (Miller et al., 2014) is a comprehensive database providing information on gene expression patterns in different brain areas throughout diverse stages of development. Gene expression levels were assessed in specific tissue samples obtained from postmortem brains from 8 weeks post-conception to 40 years of age. These ages were categorised into five distinct life stages: fetus (8–37 post-conception weeks), infant (4 months–1 year), child (2–8 years), adolescent (11–19 years), and adult (21–40 years).

### GTEx

The Genotype-Tissue Expression (GTEx) initiative represents an ongoing project to curate a publicly accessible and comprehensive dataset focused on tissue-specific gene expression patterns. Within this dataset, a remarkable compilation of 17,382 samples has been meticulously sourced from a diverse array of 53 tissue sites in 948 healthy donors. Tissue-specific gene expressions are quantified as reads per kilobase per million (RPKM). To identify differentially expressed genes (DEG) for all the tissues, log2(RPKM+1) was used. Two-sided t-tests were executed for each gene and tissue compared with all other tissues. After applying the Bonferroni correction to mitigate multiple testing issues, genes exhibiting corrected P-values below 0.05 and an absolute log fold change of at least 0.58 were classified as part of a DEG specific to a given tissue.

### MAGMA

MAGMA consists of three main steps: (1) Annotation Step: A pre-processing step is performed where GWAS-driven SNPs are mapped to genes. We employed gene locations for protein-coding genes (using Entrez gene IDs) as provided on the MAGMA website for builds 37 (hg19). SNPs located near a gene but outside its transcription region can also be mapped to that gene by specifying an annotation window around the genes. Then, we extended gene boundaries by 35 kilobase pairs (kb) upstream and 10 kb downstream. (2) Gene Analysis Step: gene p-values and other gene-level metrics were computed in this step. Correlations between neighbouring genes were also calculated in preparation for the gene-level analysis. The European panel of the 1,000 Genomes phase 3 dataset is widely utilised to address linkage disequilibrium. We selected the snp-wise=mean model for its heightened sensitivity to the mean SNP associations. (3) Gene-level Analysis: MAGMA enjoys a gene-level linear regression model. Gene sets are represented as binary indicator variables, coded as 1 for genes in the set and 0 otherwise. The association that a gene has with the phenotype is quantified as a Z-score, which is a probit transformation of the gene p-value computed during the gene analysis step (*Zg* = Φ (1 − *Pg*), mapping low p-values onto high positive Z-scores; *Zg* = 0 corresponds to *Pg* = 0.5). The competitive gene-set analysis is implemented as a linear regression model on this gene-level data matrix, *Z* = *β*0 + *Sβs* + *Cβc* + *ε*, with *S* as the gene-set indicator variable and *C* a matrix of covariates for correction. The residuals *ε* are modelled as gene-gene correlations computed during the gene analysis. The gene-set p-value results from a test on the coefficient *βs*, testing the null hypothesis *H*0: *βs* = 0 against the one-sided alternative *HA*: *βs* > 0. This tests whether the (conditional) mean association with the phenotype of genes in the gene set is higher than that of genes not in the gene set. By default, the gene set variable is conditioned on the gene size, gene density, inverse of the mean MAC in the gene and the logarithmic values of these three variables (de Leeuw et al., 2015).

## Data availability

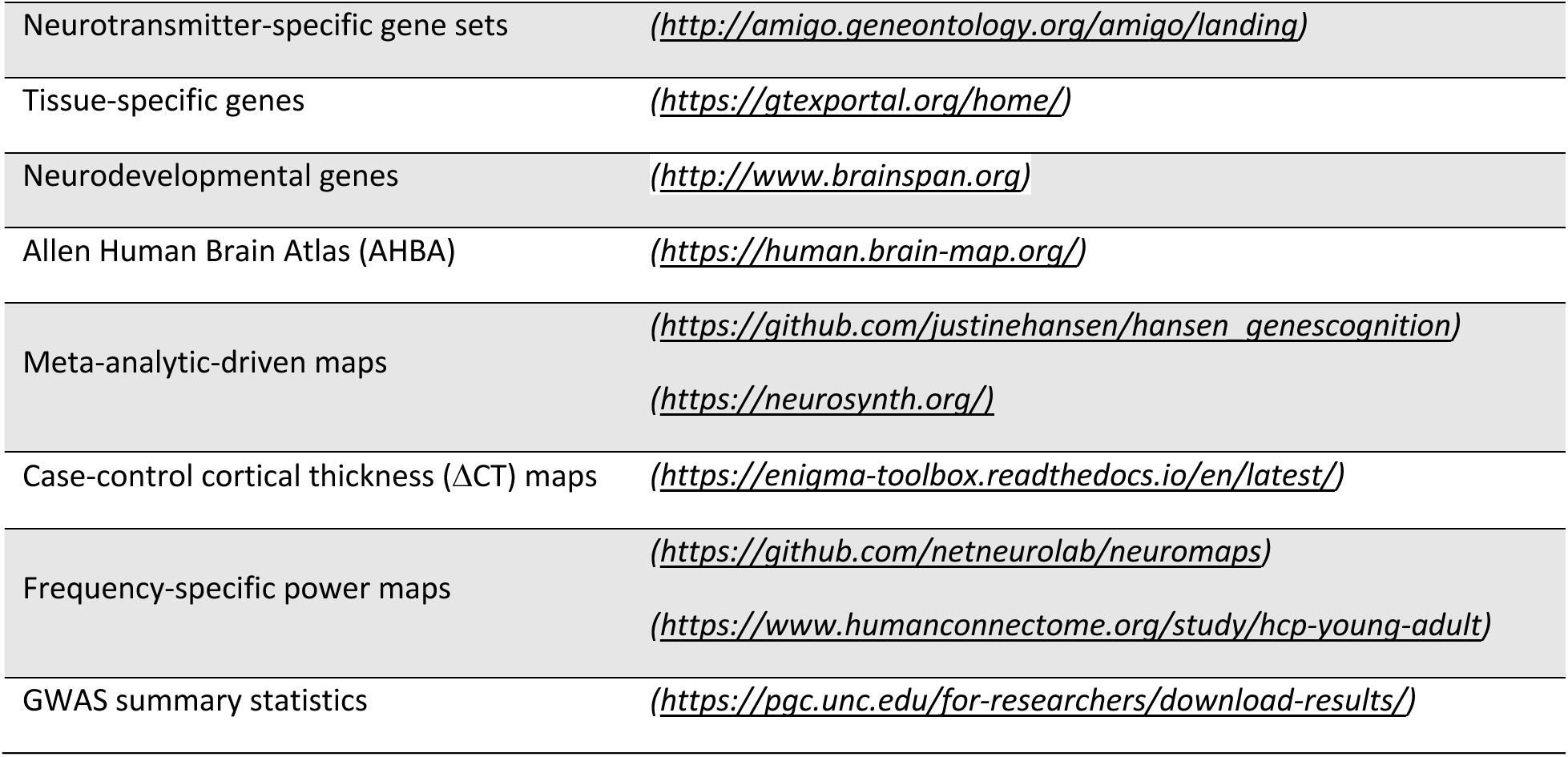

### Code, Software, or tools

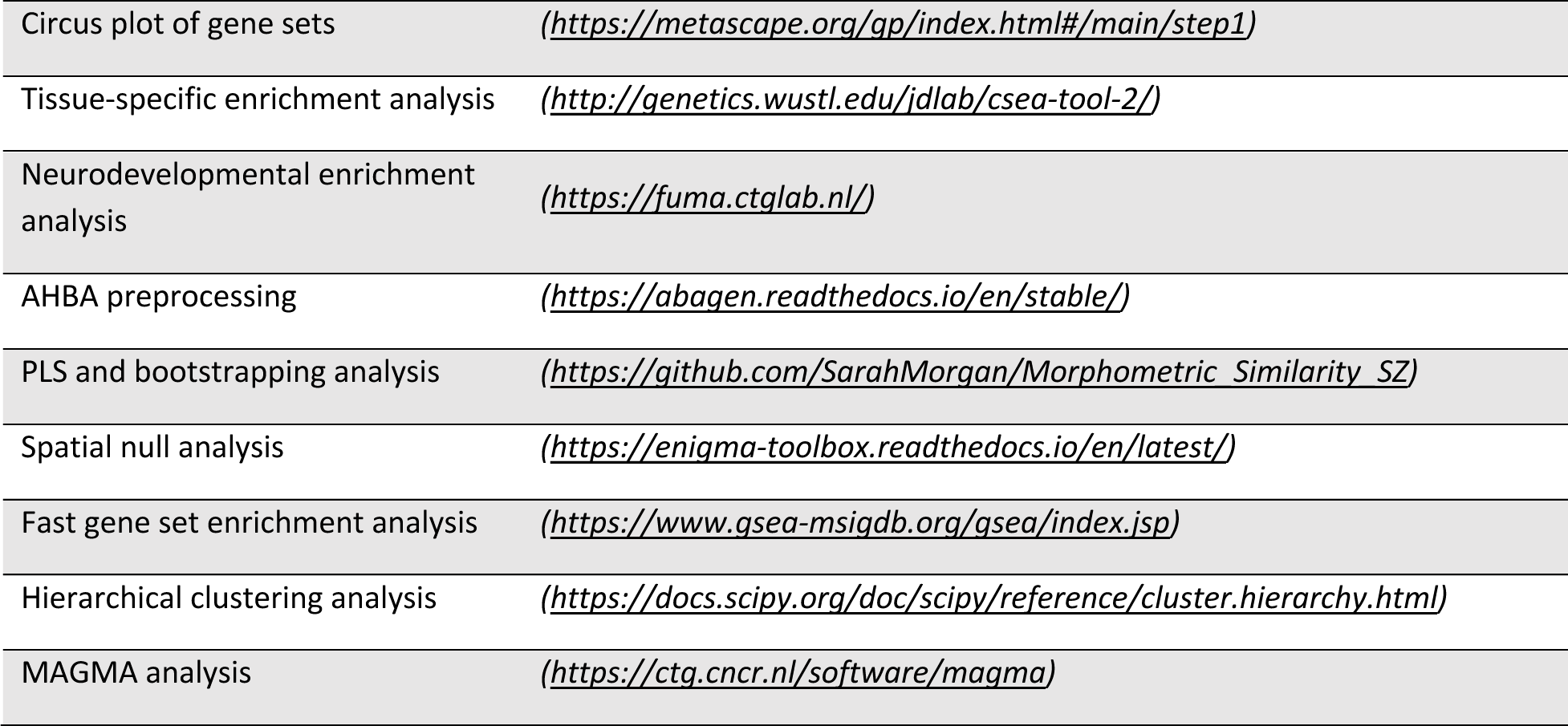

## Data Availability

All data produced in the present work are contained in the manuscript

---

**Supplementary Figure 1.**
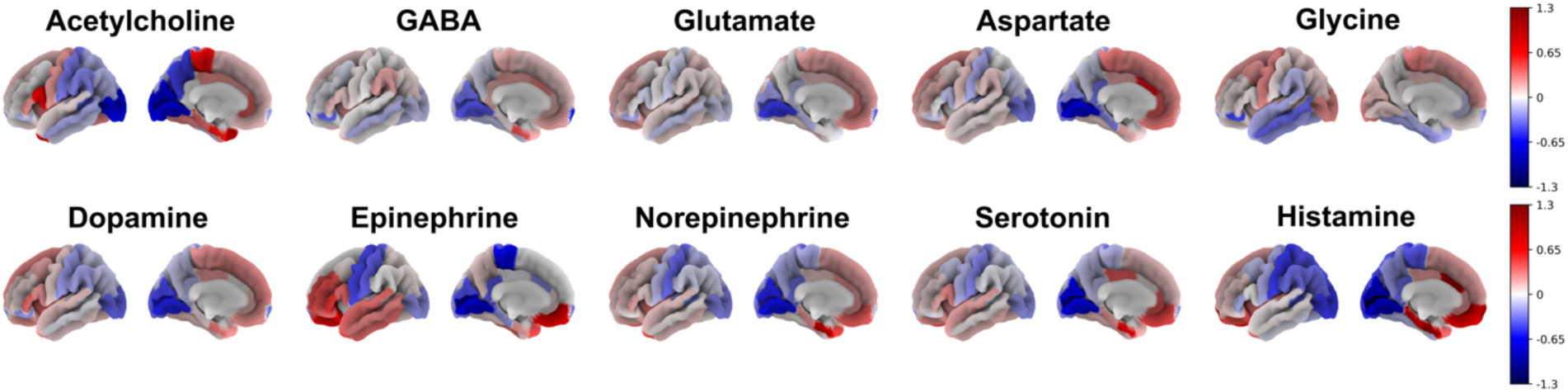
Whole-brain expression of neurotransmitter genes. The Allen Human Brain Atlas (AHBA) was employed to Investigate the neurotransmitter pathways throughout the brain. This invaluable data encompasses a comprehensive RNA microarray survey conducted on six post-mortem brains, providing gene expression profiles that are aligned with standard brain space (MNI). The transcriptional activity of 12,668 genes was mapped to the Desikan-Killiany atlas (DK). To visualise the patterns in the brain, average transcriptional values within each neurotransmitter gene set were calculated.

**Supplementary Figure 2.**
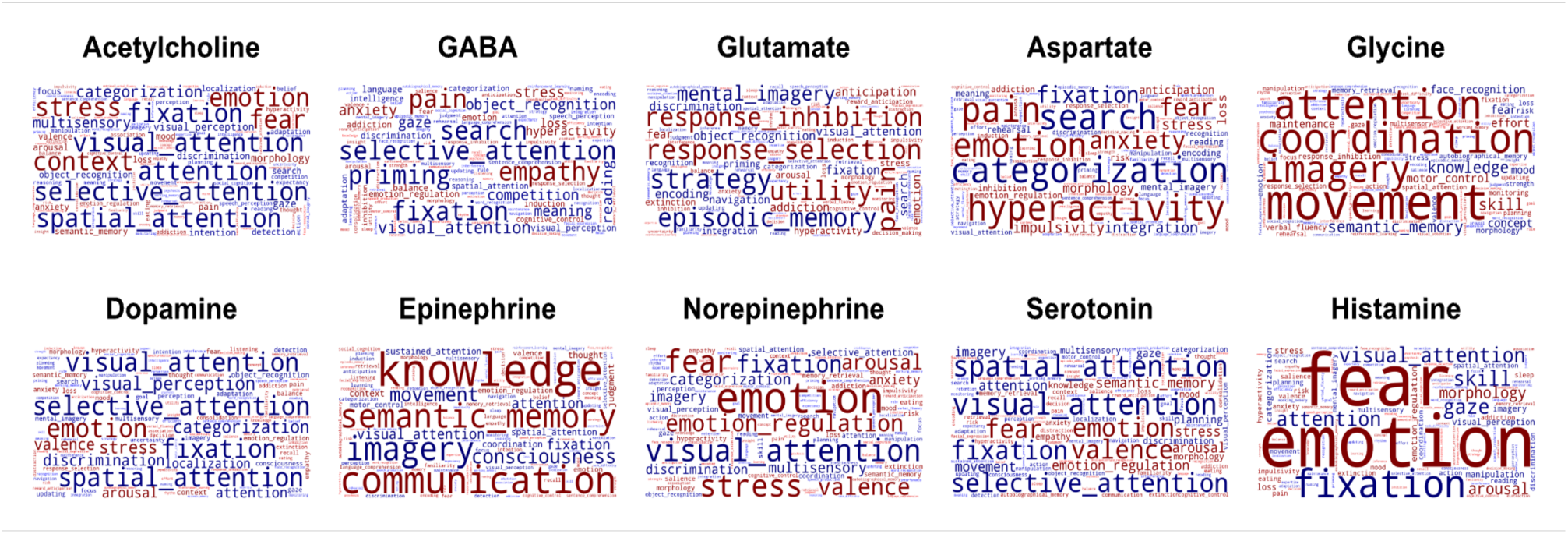
Neuropsychological translation of neurotransmitter maps. Spearman correlations were conducted between neurotransmitter maps and term-based meta-analytic-driven spatial phenotypes sourced from the Neurosynth database. A total of 123 terms from the publicly available Cognitive Atlas, a cognitive science ontology, were examined. Positive and negative correlations were indicated by red and blue colours, respectively.

**Supplementary Figure 3.**
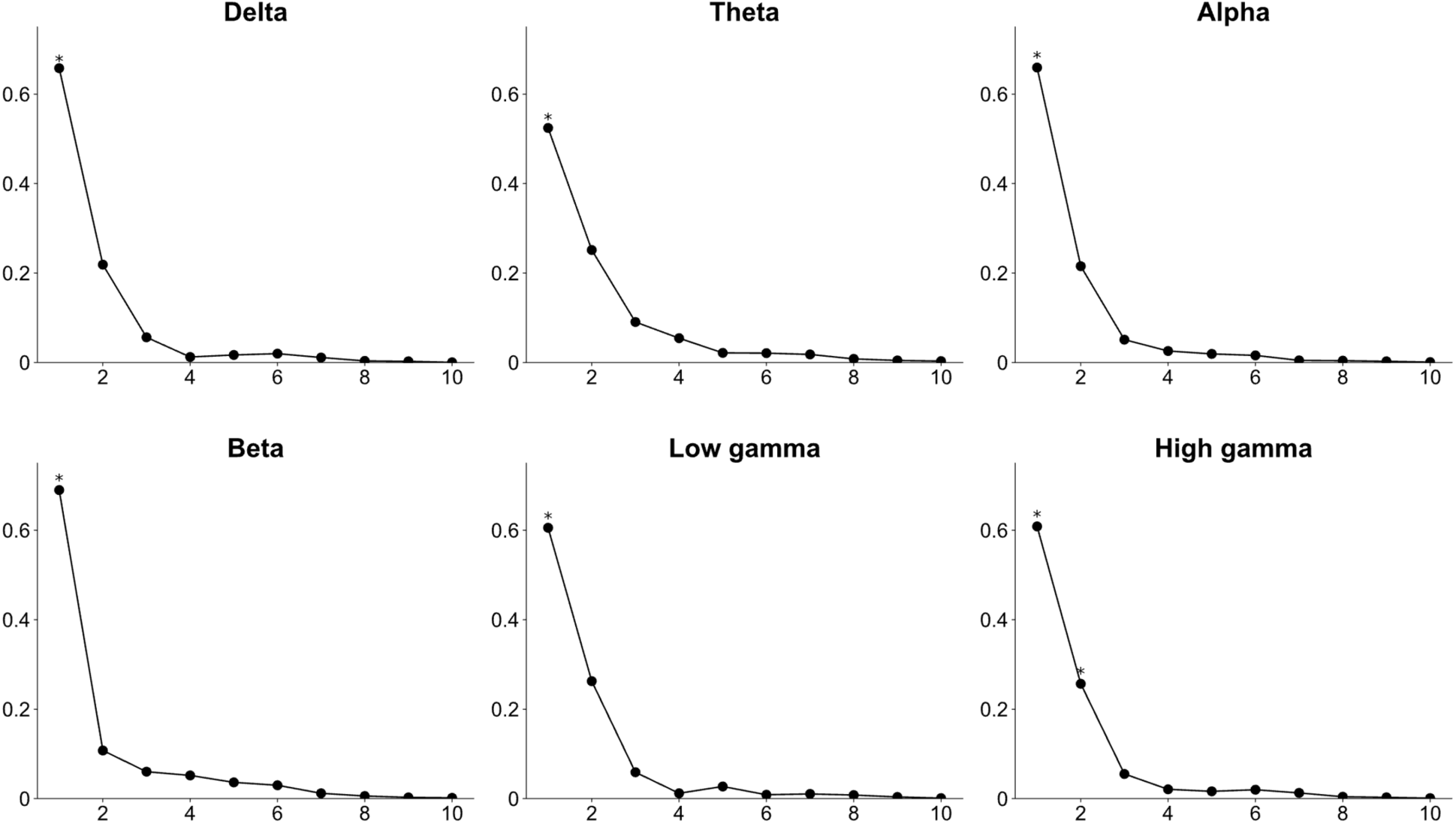
Explained variance of PLS components in transcriptional correlates of power maps. A partial least squares (PLS) analysis with brain region bootstrapping and spin permutation was performed for each power map. PLS found components from the gene expression matrix (34 regions × 12,668 genes) that have maximal covariance with each power map (34 regions × 1). PLS1 weighted maps were significantly correlated with power maps (p spin-FDR values were all < 0.01). We did not include the PLS2 component in subsequent analyses since they were not significant in all power maps and hindered us from comparing the transcriptional foundation of different power maps.

**Supplementary Figure 4.**
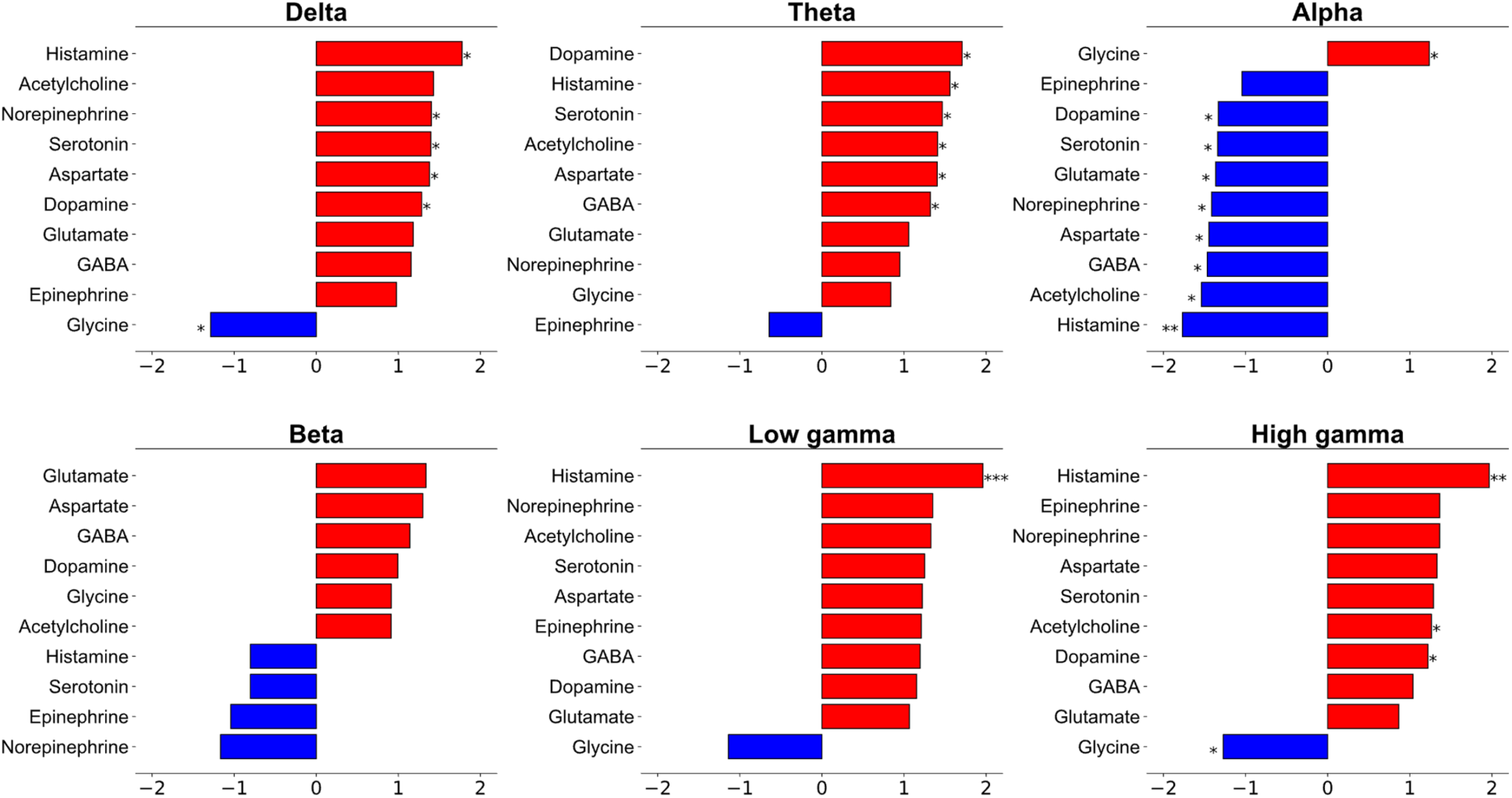
Normalised enrichment scores of neurotransmitter genes in transcriptional correlates of power maps. For each power map, we aimed to identify the positions of neurotransmitter-specific gene sets in the ranked gene list generated from the partial least square (PLS) analysis. The neurotransmitter-specific normalised enrichment scores (NES) were driven by the fast gene set enrichment analysis (FGSEA). Founders of FGSEA recommended an FDR-corrected p-value cutoff of less than 0.25. P-values are marked with one (< 0.25), two (< 0.05) and three (< 0.01) asterisks.

**Supplementary Figure 5.**
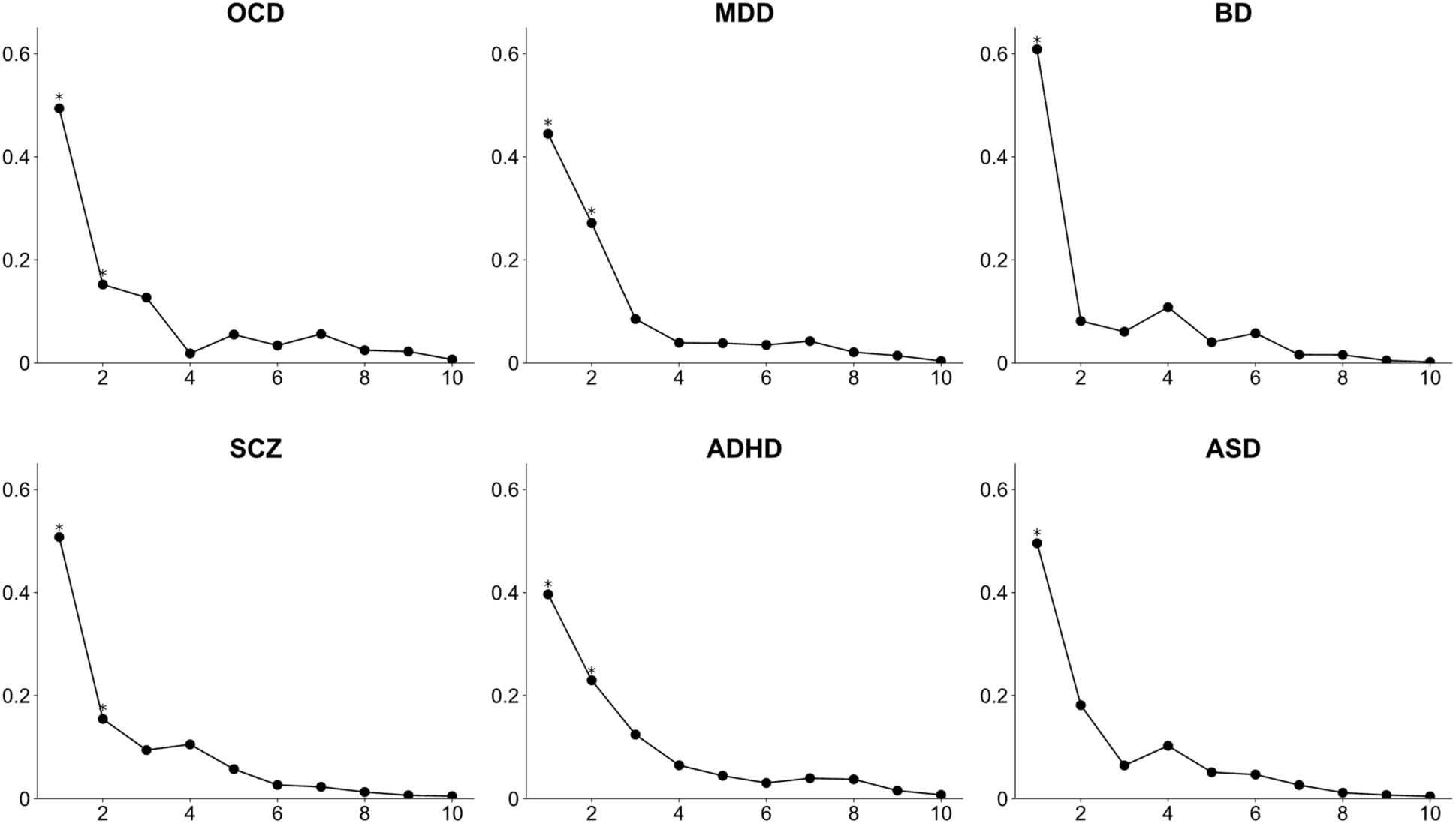
Explained variance of PLS components in transcriptional correlates of case-control cortical thickness (CT). A partial least squares (PLS) analysis with brain region bootstrapping and spin permutation was performed for each CT map. PLS found components from the gene expression matrix (34 regions × 12,668 genes) that have maximal covariance with each CT map (34 regions × 1). PLS1 weighted maps were significantly correlated with CT maps (p spin-FDR values were all < 0.01). We did not include the PLS2 component in subsequent analyses since they were not significant in all CT maps and hindered us from comparing the transcriptional foundation of different disorder-specific CT maps.

**Supplementary Figure 6.**
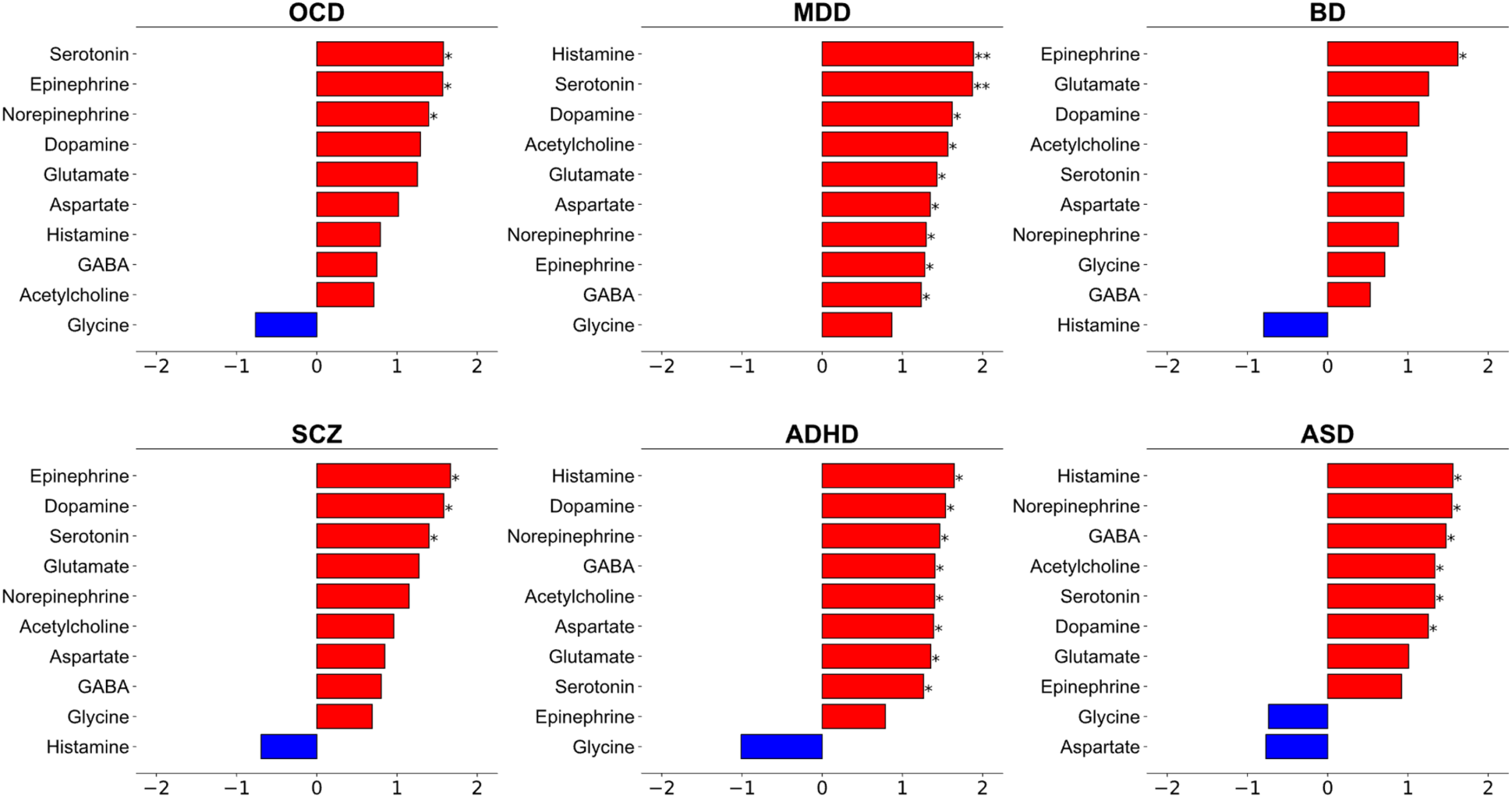
Normalised enrichment scores of neurotransmitter genes in transcriptional correlates of case-control cortical thickness (CT). For each (CT) map, we aimed to identify the positions of neurotransmitter-specific gene sets in the ranked gene list generated from the partial least square (PLS) analysis. The neurotransmitter-specific normalised enrichment scores (NES) were driven by the fast gene set enrichment analysis (FGSEA). Founders of FGSEA recommended an FDR-corrected p-value cutoff of less than 0.25. P-values are marked with one (< 0.25), two (< 0.05) and three (< 0.01) asterisks.

## Notes

### Competing Interest Statement

The authors have declared no competing interest.

### Funding Statement

This study did not receive any funding

### Author Declarations

Data was readily accessible prior to the initiation of the study, and we shared download links for easy retrieval. Neurotransmitter-specific gene sets(http://amigo.geneontology.org/amigo/landing) Tissue-specific genes(https://gtexportal.org/home/) Neurodevelopmental genes(http://www.brainspan.org) Allen Human Brain Atlas (AHBA)(https://human.brain-map.org/) Neuropsychological decoding (https://neurosynth.org/) Case-control cortical thickness maps(https://enigma-toolbox.readthedocs.io/en/latest/) Frequency-specific power maps(https://github.com/netneurolab/neuromaps) (https://www.humanconnectome.org/study/hcp-young-adult) GWAS summary statistics(https://pgc.unc.edu/for-researchers/download-results/)

